# Neurology Training in Italy: A Survey on Italian Residency Programmes and Compliance with European Training Standards

**DOI:** 10.1101/2025.02.15.25322325

**Authors:** Matteo Farè, Luca Angelini, Andi Nuredini, Bruna Nucera, Giulia Fiume, Alessandro Bombaci, Sandy Maria Cartella, SIgN

## Abstract

**Background:** Since 2005, the European Union of Medical Specialists – Section of Neurology and the European Board of Neurology have outlined a comprehensive curriculum, the European Training Requirements in Neurology (ETRN), which defines the essential clinical knowledge, practical skills, and research competencies that European neurology training programmes should encompass. Despite these standards, notable differences in training quality and resources persist among Italian residency programmes. This survey, conducted by the Italian Section of Young Neurologists, aimed to assess the alignment of Italian neurology residency programmes with ETRN standards and to evaluate residents’ and early-career neurologists’ self-reported knowledge, practical skills, and access to specialised training opportunities.

**Methods:** An anonymous, online survey was distributed to neurology residents and recently qualified neurologists across Italy in November 2023. The survey included 39 items across four sections: demographics, residency programme structure, learning objectives, and out-of-network internship opportunities. Responses were evaluated using descriptive statistics and comparative analyses across institution sizes and training years.

**Results:** Of 248 respondents (45.2% female, mean age 28.9 years), only 70.5% of ETRN’s basic training objectives were met, with lower compliance observed for advanced competencies. Practical skills like lumbar puncture and history-taking were widely acquired, but advanced diagnostic techniques (e.g., EMG, neuro-sonography) and subspecialty training (e.g., neuro-oncology, palliative care) showed limited availability. Out-of-network internship participation was low (21.7%), mainly due to institutional constraints and lack of financial support, limiting exposure to diverse clinical environments.

**Conclusions:** Italian neurology training programmes align reasonably well with ETRN standards at the basic level, though significant gaps remain in advanced training, diagnostic techniques, and interdisciplinary skills. Enhanced standardisation, improved funding for extra-network internships, and focused efforts on advanced training are essential to elevate Italian neurology residency programmes to meet European standards more fully.

## INTRODUCTION

Neurology emerged as a distinct medical specialty in England and in France in the latter half of the 19th century with the first academic chairs established in Moscow (A. Kozhevnikov, 1869) and Paris (J.M. Charcot, 1882)^1^. By the time of the first World Neurology Meeting in Bern in 1931, only a handful of European countries, including Bulgaria, Estonia, Romania, Russia, and Norway, had incorporated neurology into their medical curricula. Consequently, neurology became an independent specialty in many European nations only in the last few decades, leading to the creation of structured residency programmes^1,2^.

In Italy, neurology was recognised as an independent specialty in 1977. Prior to 1991, medical specialty training was largely unstructured and required only a period of practical experience. In 1991, a formal admission test and curriculum were introduced, with successful candidates receiving scholarships for four years of theoretical and practical training^3^. This period was extended to five years in 1996 and later reduced back to four years in 2017 by the Italian Ministry of Education, University and Research. During this time, a set of core competencies and procedural requirements was introduced, including a minimum number of practical activities to be performed, such as lumbar punctures or electrodiagnostic tests^3–5^. Despite these regulatory frameworks, previous studies revealed significant heterogeneity among neurology training programmes across Italy^3,4,6^.

To address such discrepancies and standardise training across Europe, the European Union of Medical Specialists – Section of Neurology and the European Board of Neurology (UEMS-SN/EBN) published the first European Training Requirements in Neurology (ETRN) in 2005, subsequently updated in 2016^7^ and in 2022^8^. The ETRN outlines a comprehensive curriculum encompassing clinical knowledge, practical skills, and research competencies, ensuring that neurologists are adequately prepared to address both common neurological disorders and emerging advancements in the field. According to the ETRN, neurology residency should last at least four years, consisting of two years of general clinical neurology followed by two years of advanced training in neurophysiology or subspecialties. An additional year of specialised training is also recommended^8^. Despite these requirements, postgraduate specialty training in neurology varies significantly across European countries in terms of access systems, duration, formal curriculum, training methods, evaluation tools, external rotations, and international opportunities^9,10^.

Given this context, our study aimed to evaluate the perspective of residents and young neurologists towards their training organization, to determine their subjective preparation level on various neurological topics, both theoretical and practical, and to assess the use of teaching and evaluation tools in Italian residency programmes. These results will be compared with the standards set out in the ETRN to evaluate the alignment of Italian training with European requirements. The availability of international internships and out-of-network training opportunities within Italian residency programmes was also evaluated. Standardised training is increasingly important in a globalised Europe, where the mobility of specialists offers critical opportunities for knowledge exchange and career development.

## MATERIALS AND METHODS

### Study design

The survey was created using Google Forms, in compliance with the Checklist for Reporting Results of Internet E-Surveys (CHERRIES) (***Supplementary material, File B_JMIG_CHERRIES***)^11^. The survey comprised a total of 39 items, divided into 4 sections as follows: *Demographics*, which collected data on age, sex, residency year, and residency school, with schools classified into “small,” “medium,” and “large” based on scholarships awarded over the past six years; *Residency Programme Organization*, assessing the implementation of ETRN guidelines - including basic and advanced training, rotations, tutors, and subspecialties - with responses rated on a Likert scale; *Learning Objectives*, where participants evaluated their theoretical and practical proficiency in key ETRN skills and knowledge areas; and *Out-of-Network Internships*, gathering information on national and international internship opportunities, including duration, procedures, and participants’ opinions ***Supplementary material, File A_Survey***).

For the section on learning objectives, these were organised into the three main domains outlined in the ETRN: neurological topics (question 24 of the survey, ***Table 1***), diagnostic techniques (question 25 of the survey, ***Table 1***), and practical skills (question 26 of the survey, ***Table 1***). In each domain, the primary categories from the ETRN were distilled into one or two survey items, excluding less prevalent or overly specialised objectives. Specifically, 23 items were selected for neurological topics, 11 for diagnostic techniques, and 7 for practical skills.

**Table 1.**
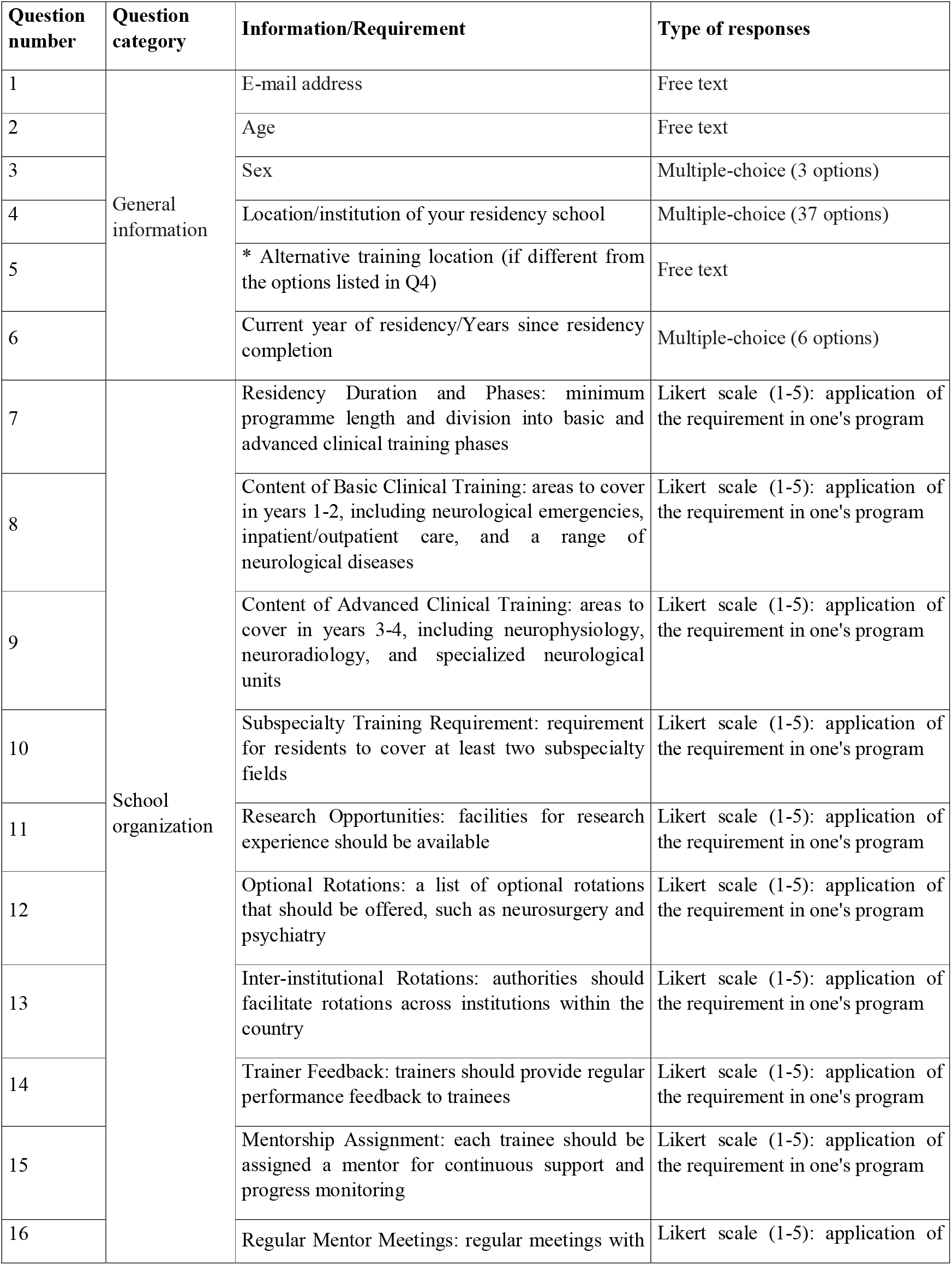
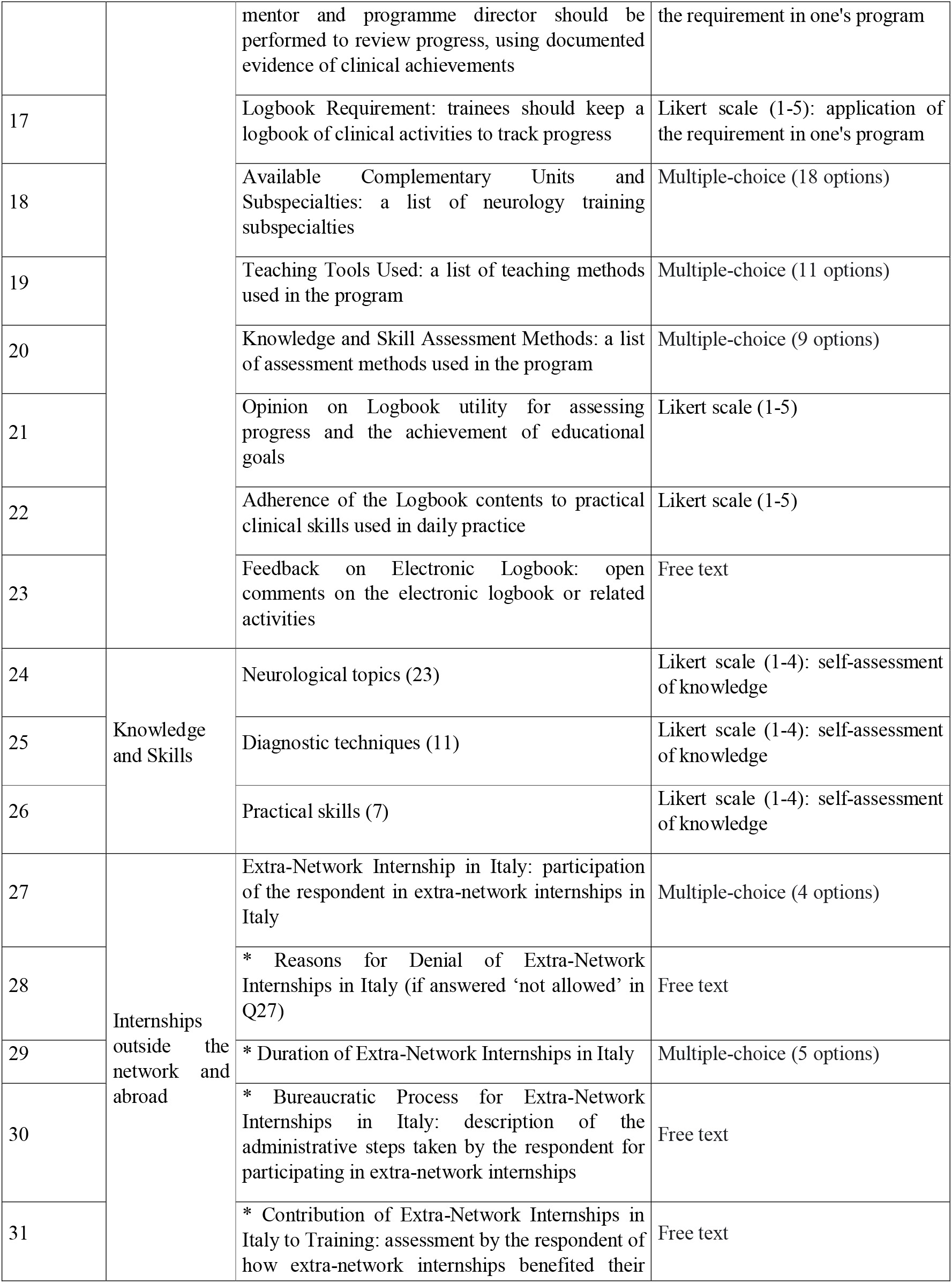
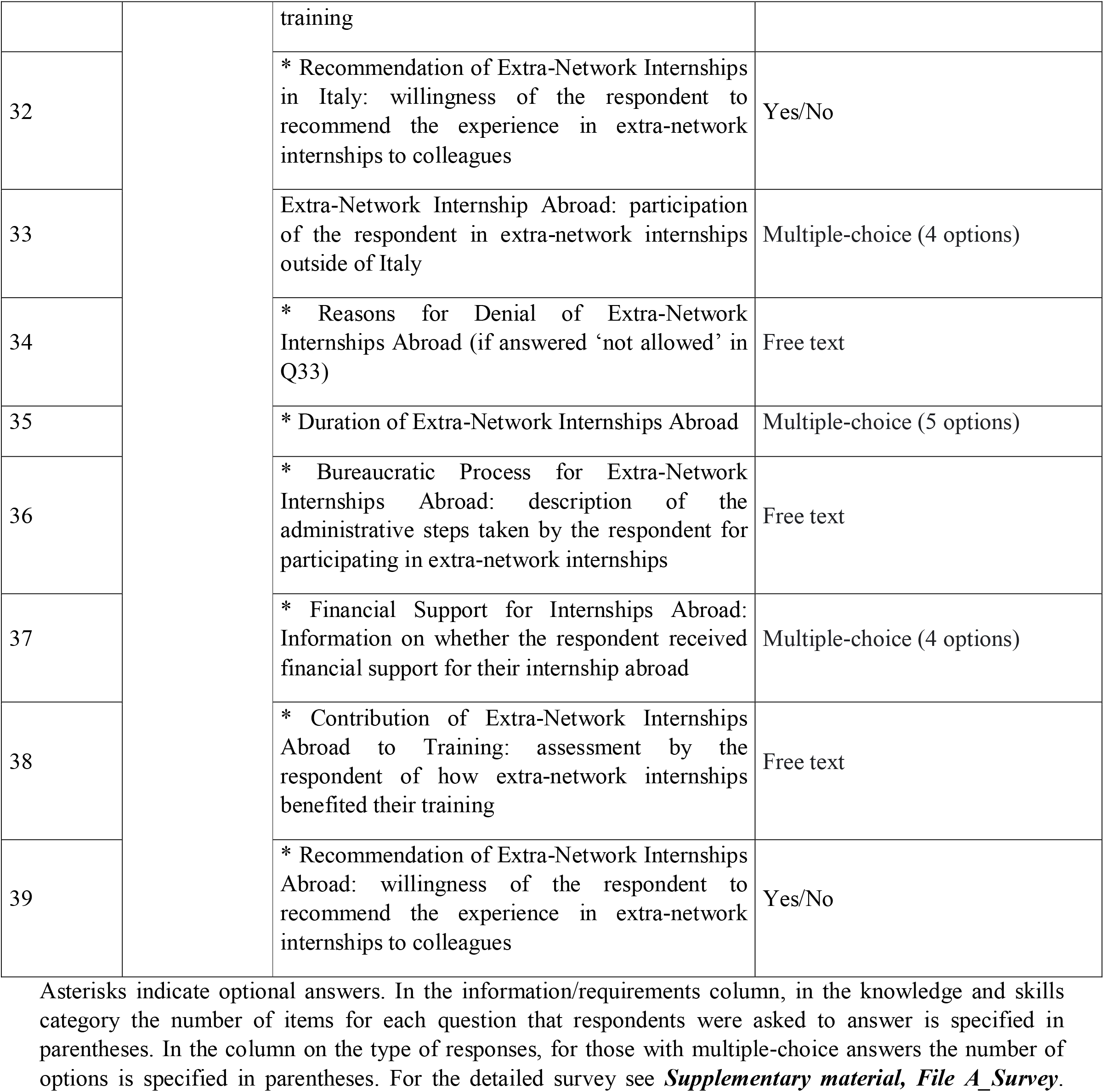
Question categories, information, and types of responses contained in the survey.

Response formats included yes/no questions, Likert scales, multiple-choice options, and free-text fields. The estimated time for completion was approximately 15 minutes. Respondents had the option to review and edit their answers before submission. No incentives were offered for participation.

The inclusion criteria required responses from Italian neurology residents and recently qualified neurologists under the age of 40. In November 2023, we contacted all potential participants directly, providing an overview of the survey’s purpose, ensuring its anonymity, stating the estimated time for completion, and including a survey link. To foster honest participation, the survey was entirely anonymous and restricted to a single submission per institutional email address to prevent duplicate responses. Personal email accounts were not permitted, and no personal information was collected. To further encourage participation, we then contacted residency programme representatives - one for each of the 36 active postgraduate neurology residency programs in Italian universities, as reported by Di Lorenzo et al.^12^ - to promote the survey among residents and neurologists under 40 who trained in their programs. Two reminders were sent by the resident representatives 15 and 30 days after the first email, and the survey was closed after two months. Ethical approval was not sought as no sensitive data were involved.

### Statistical Analysis

Descriptive statistics were calculated for all questions based on a numerical Likert scale. Responses to multiple-choice questions were reported as percentages, both overall and by school size and residency year. Shapiro-Wilk test was applied for assessing the normality of the data distribution.

For subgroup analyses, overall and pairwise comparisons were conducted using either ANOVA or the Kruskal-Wallis test, depending on the distribution of the data. The threshold for statistical significance was set at p < 0.05. All analyses were performed with Jamovi version 2.3.28.0 (the jamovi project, Sydney, Australia).

## RESULTS

### Demographic features

A total of 248 responses were obtained. Of these, 112 (45.2%) respondents identified as female, 136 (54.8%) as male, with no participants identifying as other or choosing not to disclose gender. The mean age of respondents was 28.9±2.4 years. Based on residency year, respondents were distributed as follows: 50 (20.2%) were first-year residents, 77 (31%) second-year residents, 52 (21%) third-year residents, 34 (13.7%) fourth-year residents, and 35 (14.1%) were neurologists under 40 years of age. Excluding the neurologist subgroup, the remaining 213 participants accounted for 15.1% of all neurology trainees in Italy at the time of the survey. Out of 36 neurology residency programmes in Italy, three did not respond: one small school and two medium-sized. By programme size, 93 (37.5%) respondents were from small schools, 102 (41.1%) from medium-sized, and 53 (21.4%) from large schools. Across the residency programmes, response rates varied from 1.5% to 41.7%, with a median response rate of 18.4%.

### Residency programme organization

Responses to questions on programme organisation are summarised in ***Table 2***. Higher mean scores (greater than 3) were observed for items on the structure of training into foundational and advanced phases, access to subspecialties, research facilities, and external internship opportunities within Italy. Notably, the item addressing availability of internships in external centres scored highest in this section (3.60±1.20). Lower mean scores (below 3) were recorded for questions on rotation opportunities in specific units, tutor engagement, and assessment methods, with the lowest score linked to alignment of the logbook with daily clinical practice (1.96±1.06).

**Table 2.**
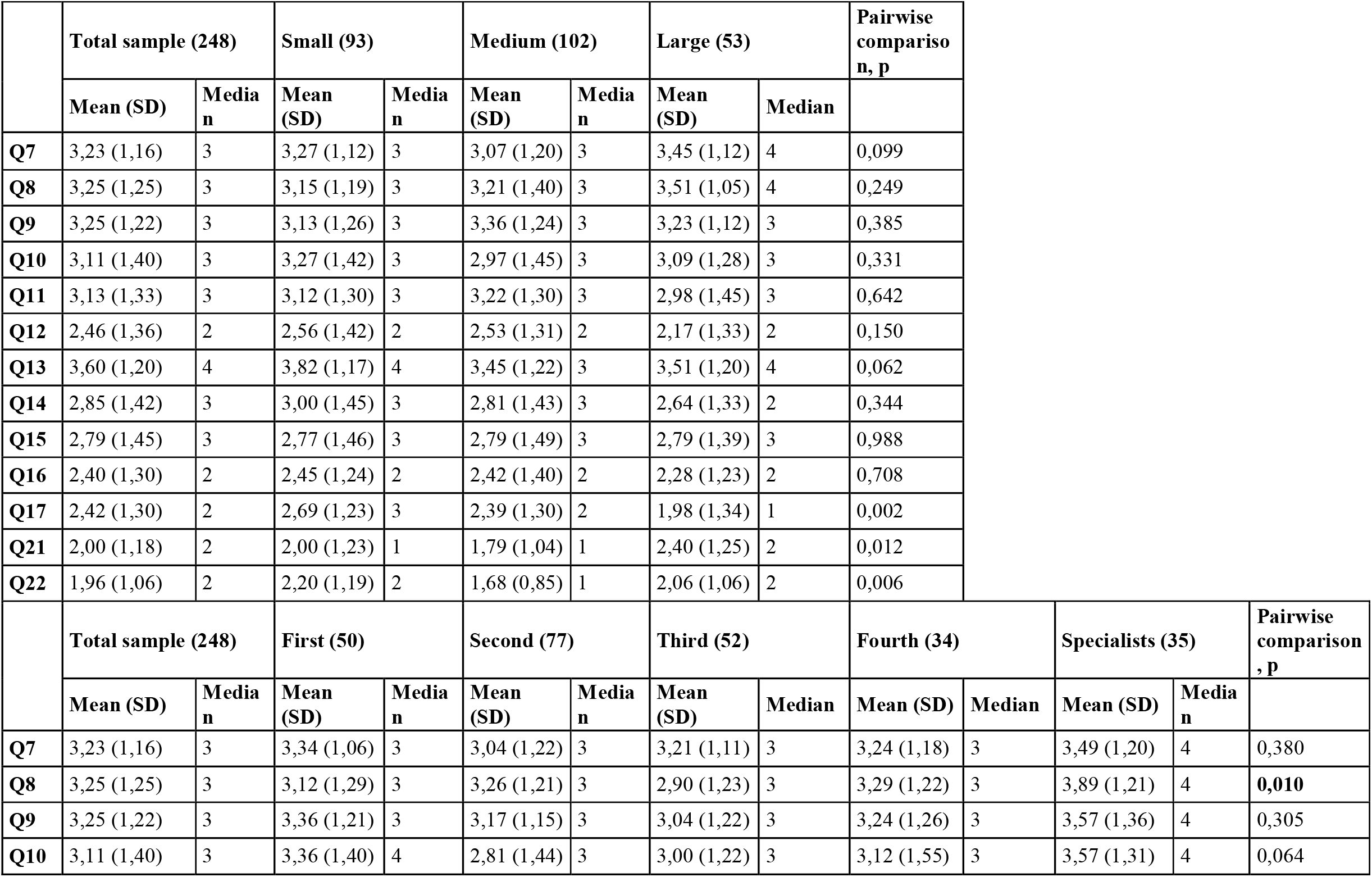
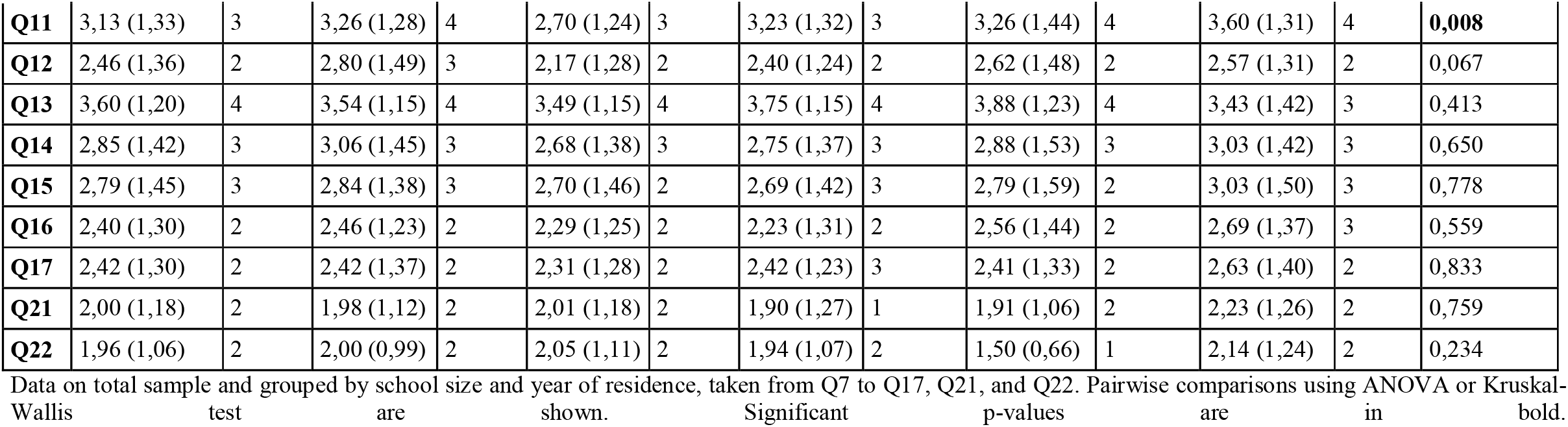
Data from questions on residency programme organization (Q7-Q17; Q21-Q22), grouped by size and year.

Analysis by programme size indicated significant differences only for logbook-related questions (Q17, Q21, Q22). Pairwise comparisons (***Supplementary material, Table 3***) showed that small programmes rated logbook use and assessment relevance higher than medium and large programmes, while large programmes scored slightly higher on logbook utility than medium programmes. Analysis by year of training showed minimal differences, primarily in foundational clinical training content and research access (Q8 and Q11).

Regarding subspecialties, the training offered was generally uniform across programme sizes (***Supplementary material, Table 4***), though shortages emerged in palliative care (3.1% of programmes), neurological intensive care (12.5%), neuropathology (15.6%), and neuro-oncology (25%).

For teaching methods (***Supplementary material, Table 4***), most schools reported using didactic lectures (84.4%), case presentations (71.4%), bedside teaching (62.5%), and clinical skill demonstrations (56.3%). Fewer schools included leadership and management training (6.3%), Grand Rounds (9.4%), or audits (18.8%).

Finally, assessment methods varied widely. Most schools used traditional exams (68.8%), written or oral, and direct observation for skills evaluation (43.8%). Only a few schools employed patient feedback (12.5%), discussion of diagnostic or therapeutic errors (15.6%), and annual review of training logbooks (18.8%), despite its formal requirement.

### Learning objectives

#### Neurological topics

The responses assessing self-reported knowledge of clinical neurological conditions indicated an overall ‘intermediate’ level of theoretical understanding (see ***Figure 1***). The majority of conditions yielded average scores between 2 (‘Knows basic concepts’) and 3 (‘Knows generally, able to make a complete diagnosis’). Only ischaemic and haemorrhagic stroke reached mean scores above 3 (3.05±0.85 and 2.92±0.88, respectively), while more common conditions scored slightly lower. Examples include Parkinson’s disease (2.57±0.92), migraine and tension-type headache (2.53±0.97 and 2.49±0.98, respectively), multiple sclerosis (2.49±0.90), epilepsy (2.47±0.89), common dementias (2.45±0.90), viral encephalitis (2.39±0.90), and polyneuropathies (2.38±0.92).

**Figure 1.**
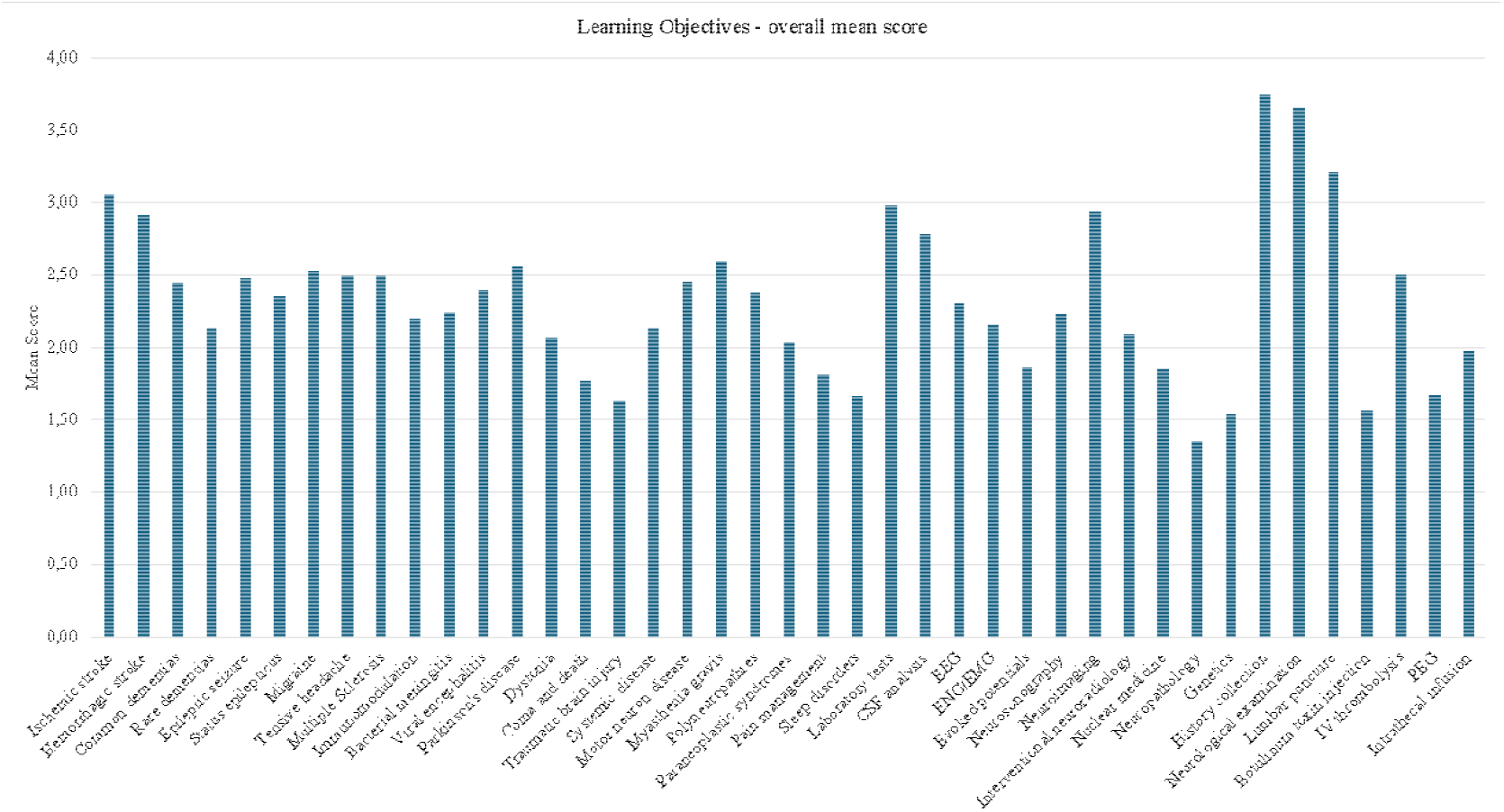
Overall mean score of learning objectives (neurological topics, diagnostic techniques, practical skills).

Lower mean scores were observed for rarer disorders or those less frequently encountered in clinical practice, such as paraneoplastic syndromes (2.04±0.87), dystonia (2.07±0.89), systemic diseases (2.13±0.78), and rare dementias (2.13±0.88). Surprisingly, certain common conditions received lower scores, sometimes even lower than rare disorders. These included pain management (1.81±0.79), coma (1.78±0.81), sleep disorders (1.67±0.81), and traumatic brain injury (1.64±0.75).

When analysed by programme size (see ***Supplementary materials, Figure 3*.*1***), average scores varied only minimally. ANOVA testing identified significant differences only for coma (p=0.02), amyotrophic lateral sclerosis (ALS) (p=0.03), and sleep disorders (p<0.01), specifically between medium and large schools (coma, p=0.03), small and medium schools (ALS, p=0.03), and small and large schools (sleep disorders, p<0.01). Analysis by residency year (see ***Figure 2***) showed a progressive increase in mean scores from first to fourth year for all topics, highlighting gradual theoretical knowledge acquisition throughout the training programme.

**Figure 2.**
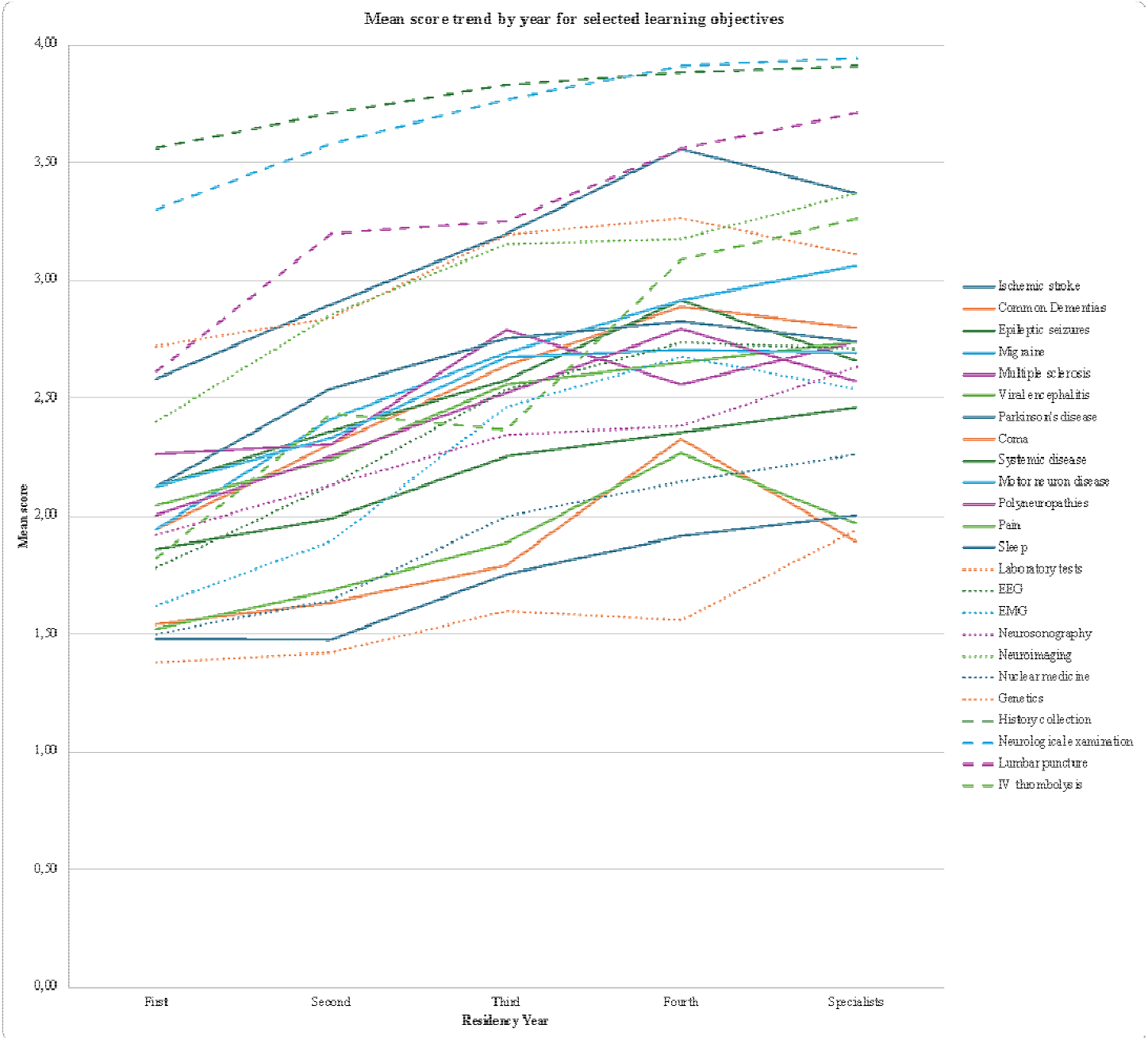
Selected learning objective, time trend by residency year.

#### Diagnostic techniques and practical skills

The data on diagnostic techniques and practical skills reflected a moderate level of confidence, with average scores between 2 (‘Can manage with assistance’) and 3 (‘Can generally manage but may need assistance’). Consistent with findings in neurological topics, no significant score differences were seen across programme sizes (see ***Supplementary materials, Figures 3*.*2 and 3*.*3***). Higher scores were recorded for commonly used techniques and routine skills, such as laboratory tests (2.99±0.92), neuroimaging (2.94±0.84), cerebrospinal fluid (CSF) analysis (2.79±0.97), history taking (3.76±0.46), neurological examination (3.66±0.52), lumbar puncture (3.22±0.80), and intravenous thrombolysis (2.50±1.05). In contrast, more specialised techniques and advanced skills had lower scores, including interventional neuroradiology (2.10±0.94), evoked potentials (1.87±0.98), nuclear medicine (1.85±0.89), genetics (1.54±0.80), neuropathology (1.36±0.63), intrathecal infusion (1.98±1.06), percutaneous endoscopic gastrostomy management (1.68±0.85), and botulinum toxin injection (1.57±0.91). Notably, common diagnostic techniques like electroencephalography (EEG) (2.31±0.97), electroneuromyography (EMG) (2.16±1.04), and neuro-sonography (2.24±1.03) also received low scores, suggesting potential training gaps.

Unlike theoretical knowledge, school size influenced scores in diagnostic methods and practical skills, with larger schools generally scoring lower than smaller ones. Significant differences were found for EEG (p<0.01), evoked potentials (p <0.01), neuro-sonography (p<0.01), nuclear medicine (p<0.01), neuropathology (p=0.02), lumbar puncture (p=0.02), and botulinum toxin injection (p=0.012). Analysis by training year (see ***Figure 2***) showed a positive trend in scores from first through fourth year, with newly qualified neurologists achieving further score increases, particularly in diagnostic techniques and practical skills, suggesting continued skill development post-specialisation.

### Comparison with ETRN expected levels

The mean scores obtained by second- and fourth-year specialization cohorts were then compared with the basic and advanced target levels proposed by the ETRN (see ***Figure 4***), considering reached a Learning Objective with a mean score not inferior to 0.5 points than the ETRN expected level.

**Figure 4.**
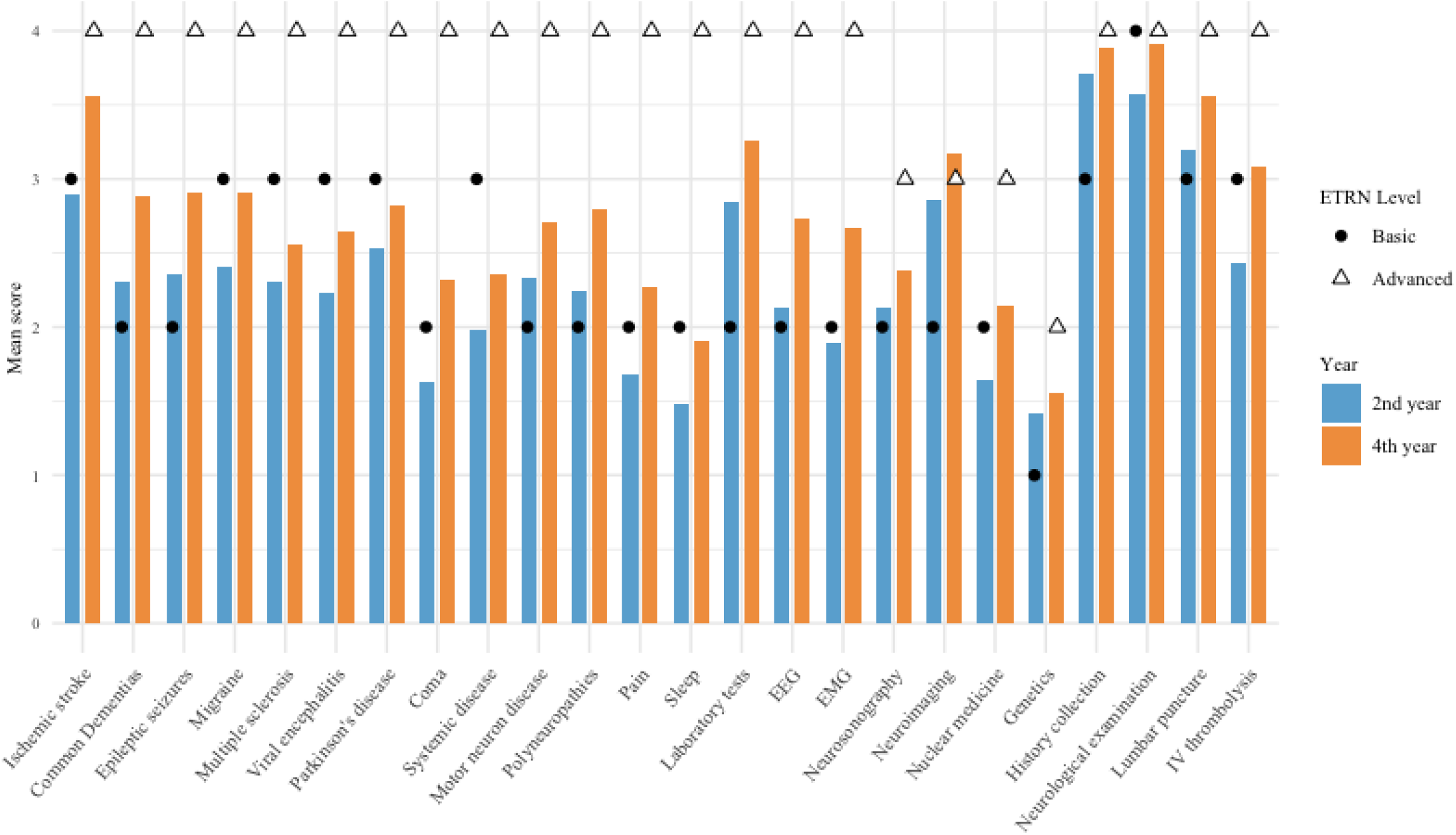
Comparison of mean score in second and fourth year of selected learning objectives with expected basic and advanced ETRN levels.

In terms of neurological topics, 14 out of 23 items (60.9%) for second-year trainees and only 2 items (8.7%) for fourth-year trainees met or exceeded the ETRN’s thresholds. For diagnostic techniques, all 11 items (100%) reached the basic level, though only 2 techniques (18.2%), specifically neuroimaging and genetics, achieved the advanced level. Regarding practical skills, 5 out of 7 items (71.4%) reached the basic level, and 4 out of 7 items (57.1%) met the advanced level, with minimal difference between second- and fourth-year scores.

Overall, these results indicate that a substantial portion of competencies reached the basic target level (30 out of 41; 73.1%), while a smaller proportion met the advanced level (8 out of 41; 19.5%). Notably, for 12 items (29.3%), neither the basic nor advanced level was achieved, and these competencies showed minimal improvement across training years, likely reflecting an underlying lack of emphasis in residency training (see ***Figure 2***).

### Out-of-network Internships

Survey findings reveal that only 54 out of 248 trainees (21.7%) participated in out-of-network internships (both in Italy and outside) during their residency, with 40 (16.1%) undertaking placements at affiliated institutions and 14 (5.6%) at non-affiliated institutions. The duration of these placements typically ranged from 3 to 9 months, with 40.0% lasting 3–6 months, 25.5% lasting 6–9 months. Shorter placements of under 3 months were reported by 27.2% of participants, and only 7.3% extending beyond 9 months.

International placements were even less common, with only 28 trainees (10.5%) gaining experience abroad, primarily in EU countries (20 trainees, or 8.1%) and less frequently in non-EU countries (8 trainees, or 2.4%). Most international placements spanned 3–6 months (35.7%) or 6–9 months (35.7%), with fewer lasting under 3 months (17.9%) or 9–12 months (10.7%). None exceeded one year. Financial support was limited: only 8 of 28 respondents (28.6%) reported receiving institutional funding, while 20 (60.7%) received no financial aid despite expressing a need for it.

Analysis by residency year and programme size showed no significant correlations, likely due to the limited sample size. Notably, 12 (4.8%) and 13 (5.2%) participants reported that their requests for domestic and international placements, respectively, were denied. Common reasons for refusal included institutional restrictions limiting these placements to final-year trainees, perceived disruption to clinical responsibilities, and doubts about the relevance of the proposed placements. Secondary barriers included the COVID-19 pandemic and lack of responses from host centres.

Despite these challenges, all trainees who completed out-of-network internships reported high satisfaction, highlighting considerable educational and professional gains. Participants unanimously recommended these experiences, endorsing them as valuable additions to residency training, both within Italy and internationally.

## DISCUSSION

This survey, undertaken by the Italian Section of Young Neurologists, aimed to evaluate the quality and scope of educational and clinical training within Italian neurology residency programmes, with a particular focus on their alignment with the ETRN standards. Rigorous monitoring of training programmes is essential to ensure consistent and high-quality preparation for future neurologists across Italy^13^.

This initiative builds upon prior research, notably a 2005 study by Facheris et al.^3^, which investigated similar aspects of neurology residency training in Italy and a previous survey conducted by Bonifati et al.^6^. Both studies had already unveiled a significant variability in neurology training in Italy, attributing in part the differences to the lack of internal or external monitoring and local healthcare problems. These issues have been partially addressed by the implementation of the Osservatorio Nazionale della Formazione Medica Specialistica, that was established by the Legislative Decree No. 368 of August 17, 1999 (Article 43) and later amendments to set training standards and monitor the quality of specialist medical education in Italy. A European survey conducted by Kleineberg et al.^9^, under the auspices of the Resident and Research Fellow Section of the European Academy of Neurology (EAN), indicated that in 80% of 32 European countries, the national Neurology Society plays an active role in shaping training programmes. In Italy, this role is assumed by the Ministry of University and Research. Moreover, application procedures differ markedly across Europe; in some countries, candidates submit applications through scientific societies, while in others, they apply directly to hospitals. Since 2017, Italy has implemented a nationwide multiple-choice examination for residency admission, standardising the entry process^14^.

Our survey captured data from 15.1% of neurology residents and early-career specialists and provides a balanced demographic representation (45.2% female, 54.8% male) across 33 of the 36 Italian neurology residency programmes. The mean response rate of 18.4% per institution ensured that the sample reflected the age, gender, residency year, and school size distributions of the larger population. A significant concern identified in this survey pertains to the organizational structure of training, particularly the ETRN’s recommendation to delineate residency into “basic” and “advanced” stages. This recommendation is only moderately implemented across Italian training programmes, receiving an average score of 3 out of 5, without notable discrepancies between larger and smaller programmes.

The survey also found low scores for optional rotations in departments outside the core neurology training, possibly due to the time constraints imposed by the comprehensive skill and knowledge requirements of the standard residency curriculum. Furthermore, mentorship meetings and logbook documentation were poorly rated, particularly in larger institutions where logbooks are less commonly used. While larger programmes reported the portfolio system as beneficial, limited implementation suggests that accessibility challenges may impact trainees’ experience in these institutions.

Focusing on the evaluation of theoretical and clinical knowledge, respondents reported significant gaps in training regarding diseases included in the ETRN core curriculum, such as dementia, epilepsy, multiple sclerosis, infectious diseases, coma, and death.

These areas received particularly low scores for perceived proficiency, suggesting that residency training in these topics may be insufficient, especially for conditions often managed in specialised centres that are not uniformly accessible to residents. In line with the findings by Bocci et al.^4^, advanced diagnostic techniques, such as EMG, evoked potentials, and cerebrovascular neuro-sonography, were also flagged as inadequately covered. While these skills are essential components of advanced neurology training per ETRN standards, they may be restricted by limited access to specialised centres and by the necessity for interprofessional collaboration, such as with interventional neuroradiologists and nuclear medicine specialists.

Moreover, topics requiring multidisciplinary collaboration—including traumatic brain injury, systemic disease, paraneoplastic syndromes, and pain management—also yielded low scores. The absence of dedicated pathways within residency programmes for these conditions highlights an area ripe for enhancement, particularly as they are encompassed within the ETRN guidelines. Specialised conditions like motor neuron disease, polyneuropathy, and sleep disorders are frequently restricted to centres with highly specific expertise, potentially limiting residents’ exposure to these critical pathologies.

Data from recently qualified neurologists suggests that training in practical skills and diagnostic techniques continues beyond residency. This continuation may indicate a need for extended education to bridge gaps identified in residency. Collecting data from more senior neurologists could provide valuable insights into the long-term effects of residency training on skill acquisition and retention.

When analysing subspecialty training, minimal differences were observed in offerings across institutions of varying sizes. However, fields such as neuro-oncology, neuropathology, neurological intensive care, and palliative care were frequently underrepresented. The specialised nature of these domains, particularly palliative care and neuro-oncology, coupled with the requirement for shared care pathways with other disciplines, likely contributes to their limited availability. Notably, palliative care education has been the focus of a recent Italian survey^15^. Limited laboratory resources and infrastructure in neuropathology were cited as additional obstacles to comprehensive training in this field.

Interestingly, the sub-analysis based on school size reveals a consistent trend: smaller residency programmes tend to achieve higher scores in practical diagnostic skills, whereas larger programmes exhibit comparatively lower scores. Pairwise analysis confirms these differences, indicating that smaller schools receive more favorable scores compared to medium and/or large institutions. This trend may suggest that larger student bodies within residency programmes dilute practical training opportunities, potentially resulting in a more fragmented learning experience. However, given the limited data from larger schools, this observation warrants further investigation to validate the impact of programme size on practical skills training.

Furthermore, doing a comparison with other European countries, training in pain management, neurotraumatology, and neuro-oncology is often constrained due to overlaps with other specialities. Neurophysiology training, too, is inconsistently provided across Europe, with only 66% of countries incorporating it into neurology residency programmes. While stroke management training in Italy is relatively well-developed, the skill of thrombectomy remains exclusive to neuroradiologists, unlike in countries such as France and Turkey, where neurologists are also trained in this procedure. While theoretical and practical instruction is generally consistent across institutions, programmes addressing leadership, project management, Grand Rounds, and audit activities remain inadequate. Evaluation methods are similarly variable, with limited integration of patient feedback, discussion of diagnostic or treatment errors, and evaluation of logbooks—core components of the ETRN framework.

Looking at learning objectives, only in 8 items out of 41 concerning ETRN, respondents indicated to have reached the advanced level, while in 12 items even the basic level was not reached.

These findings indicate the difficulty that Italian residency programmes face in fully aligning with ETRN standards. Notably, for most items, second-year scores did not meet the ETRN basic target, and little to no growth in scores was observed by the fourth year, particularly for diagnostic techniques. This trend may reflect a lack of long-term training strategies in Italian neurology residencies. In contrast, practical skills such as history-taking, examination, and lumbar puncture, which are largely acquired during the first two years, show satisfactory alignment with ETRN standards, but minimal improvement is seen in later years.

The survey also highlights significant barriers for Italian neurology trainees in accessing extra-network internships, with only 21.7% of respondents participating in such experiences, and a mere 10.5% completing placements abroad. Institutional policies and logistical challenges appear to limit these opportunities, as evidenced by reports of denied requests based on residency year, clinical responsibilities, and prioritisation of in-network training. Financial support for these internships is also limited, with most trainees expressing a need for funding, which could disproportionately impact residents from less privileged backgrounds and exacerbate disparities in training opportunities.

A notable limitation of this study is the reliance on self-assessment, which may lead residents to undervalue their competencies. This tendency to underestimate skills could result in findings that reflect perceived rather than actual proficiency. Such discrepancies might obscure strengths or improvements achieved during training, affecting the overall accuracy of the data. Future studies could incorporate objective assessments or external evaluations to provide a clearer view of residents’ true competencies. Another limitation that needs to be considered is the limited numerosity, which can limit the generalization of our findings. However, the sample size remains adequate for the study. Moreover, it is well-balanced in terms of geographic distribution, and nationwide survey access was ensured through effective dissemination of information and accessibility for all residents. Furthermore, a selection bias must be addressed, since it is possible that unsatisfied residents were more eager to take the survey, therefore leading to a decrease in overall scores. Finally, a further limitation may relate to some respondents having completed part of their training during the COVID-19 pandemic, potentially influencing both their educational experience and their self-perception as nascent neurologists. A prior study by the Italian Section of Young Neurologists^12^ underscored how the reorganisation of clinical services, marked by reduced elective admissions, curtailed outpatient follow-ups, and the introduction of telemedicine, significantly altered traditional training models. During the initial emergency phase, theoretical teaching was either cancelled or rescheduled across Italian universities. Larger studies are needed to confirm our findings.

Despite the variability in training quality, our survey highlights several strengths within Italian neurology residency programmes. Most notably, foundational training is well structured, ensuring that residents acquire core competencies in history-taking, neurological examination, and lumbar puncture early in their training. Additionally, residents who completed extra-network placements when available reported a high level of satisfaction, underlining the educational and professional value of exposure to diverse clinical environments. Furthermore, the presence of research opportunities in many programmes represents a key strength that aligns with European standards, fostering early-career engagement in academic neurology.

## CONCLUSIONS

Overall, Italy’s neurology training programmes align reasonably well with European standards, however, notable gaps persist in standardization and quality assessment across institutions. While opportunities for improvement exist, Italy is largely comparable to other European countries in most areas, except for the lack of a unified training framework and systematic quality evaluation.

To further improve neurology training in Italy, institutions should focus on standardizing the implementation of advanced training across residency programmes. Ensuring structured exposure to subspecialties such as neuro-oncology, neuropathology, and neuro-intensive care will provide residents with a more comprehensive skill set. Additionally, expanding access to out-of-network internships, both nationally and internationally, by simplifying administrative procedures and offering targeted financial support could significantly enhance training quality. Strengthening mentorship structures and logbook utilization would also provide clearer learning pathways and improve skill assessment. By addressing these areas, Italian neurology residency programmes can bridge existing gaps while capitalizing on their existing strengths to meet and exceed European training standards.

## Data Availability

Anonymized data not published within this article will be made available by request from any qualified investigator.

## Acknowledgment

We thank all the Italian young neurologists for participating in the survey. We thank the Italian Section for Young Neurologists of the Italian Society of Neurology (SIN). We acknowledge the President of the Italian Society of Neurology, Prof. Alessandro Padovani, for his support and his invaluable supervision and intellectual contribution to this work.

