## Supplementary materials for "Neurology Training in Italy: A Survey on Italian Residency Programmes and Compliance with European Training Standards"

**Table 3. Data on residency programme organization (Q7-Q17; Q21-Q22), grouped by school size; pairwise comparison (DSCF method).**

|  |  |  | **p** |  |  |  | **p** |  |  |  | **p** |
| --- | --- | --- | --- | --- | --- | --- | --- | --- | --- | --- | --- |
| **Q7** | Large | Medium | 0,098 | **Q12** | Large | Medium | 0,158 | **Q16** | Large | Medium | 0,922 |
|  | Large | Small | 0,551 |  | Large | Small | 0,206 |  | Large | Small | 0,677 |
|  | Medium | Small | 0,401 |  | Medium | Small | 0,999 |  | Medium | Small | 0,881 |
| **Q8** | Large | Medium | 0,487 | **Q13** | Large | Medium | 0,950 | **Q17** | Large | Medium | 0,072 |
|  | Large | Small | 0,174 |  | Large | Small | 0,237 |  | Large | Small | **0,002** |
|  | Medium | Small | 0,919 |  | Medium | Small | 0,064 |  | Medium | Small | 0,207 |
| **Q9** | Large | Medium | 0,663 | **Q14** | Large | Medium | 0,770 | **Q21** | Large | Medium | **0,007** |
|  | Large | Small | 0,920 |  | Large | Small | 0,323 |  | Large | Small | 0,113 |
|  | Medium | Small | 0,389 |  | Medium | Small | 0,648 |  | Medium | Small | 0,598 |
| **Q10** | Large | Medium | 0,860 | **Q15** | Large | Medium | 0,992 | **Q22** | Large | Medium | 0,088 |
|  | Large | Small | 0,687 |  | Large | Small | 0,987 |  | Large | Small | 0,822 |
|  | Medium | Small | 0,319 |  | Medium | Small | 0,999 |  | Medium | Small | **0,006** |
| **Q11** | Large | Medium | 0,639 |  |  |  |  |  |  |  |  |
|  | Large | Small | 0,862 |  |  |  |  |  |  |  |  |
|  | Medium | Small | 0,859 |  |  |  |  |  |  |  |  |

Data on residency programme organization, grouped by school size, taken from Q7 to Q17, Q21, and Q22. Pairwise comparisons using Kruskal-Wallis test are shown. Significant p-values are in bold.

**Table 4. Data on available subspecialties, teaching methods, and evaluation tools used (Q18-Q20), grouped by school size**

| **AVAILABLE SUBSPECIALTIES** | **Total, N (%)** | **Small, N (%)** | **Medium, N (%)** | **Large, N (%)** |
| --- | --- | --- | --- | --- |
| Alzheimer's and dementia | 27 (84,4) | 10 (76,9) | 12 (85,7) | 5 (100) |
| Cerebrovascular diseases | 27 (84,4) | 11 (84,6) | 11 (78,6) | 5 (100) |
| Epilepsy | 30 (93,8) | 12 (92,3) | 13 (92,9) | 5 (100) |
| Headaches | 28 (87,5) | 11 (84,6) | 12 (85,7) | 5 (100) |
| Movement disorders | 29 (90,6) | 11 (84,6) | 13 (92,9) | 5 (100) |
| Multiple Sclerosis and demyelinating diseases | 29 (90,6) | 11 (84,6) | 13 (92,9) | 5 (100) |
| Neuro-oncology | 8 (25) | 1 (7,7) | 5 (35,7) | 2 (40) |
| Neurological/neurosurgical intensive care unit | 4 (12,5) | 1 (7,7) | 1 (7,1) | 2 (40) |
| Neuromuscular diseases | 28 (87,5) | 11 (84,6) | 12 (85,7) | 5 (100) |
| Neuropathology | 5 (15,6) | 0 (0) | 4 (28,6) | 1 (20) |
| Neurophysiology | 26 (81,3) | 10 (76,9) | 11 (78,6) | 5 (100) |
| Neuroradiology | 22 (68,8) | 7 (53,8) | 11 (78,6) | 4 (80) |
| Neurorehabilitation | 13 (40,6) | 3 (23,1) | 6 (42,9) | 4 (80) |
| Palliative Care and Hospice | 1 (3,1) | 0 (0) | 1 (7,1) | 0 (0) |
| Peripheral nerve diseases | 26 (81,3) | 10 (76,9) | 11 (78,6) | 5 (100) |
| Sleep disorders | 9 (28,1) | 3 (23,1) | 6 (42,9) | 2 (40) |
| Stroke Unit | 27 (84,4) | 12 (92,3) | 10 (71,4) | 5 (100) |
| **TEACHING METHODS** | **Total, N (%)** | **Small, N (%)** | **Medium, N (%)** | **Large, N (%)** |
| Audit | 6 (18,8) | 1 (7,7) | 4 (28,6) | 1 (20) |
| Case presentations | 23 (71,9) | 8 (61,5) | 12 (85,7) | 3 (60) |
| Demonstration and teaching of clinical skills | 18 (56,3) | 4 (30,8) | 12 (85,7) | 2 (40) |
| Evidence-Based Medicine and Journal Club | 14 (43,8) | 4 (30,8) | 8 (57,1) | 2 (40) |
| Grand Rounds | 3 (9,4) | 2 (15,4) | 0 (0) | 1 (20) |
| Leadership and project management programmes | 2 (6,3) | 0 (0) | 2 (14,3) | 0 (0) |
| Lectures and/or small groups | 27 (84,4) | 11 (84,6) | 11 (78,6) | 5 (100) |
| Multi-disciplinary meetings | 12 (37,5) | 4 (30,8) | 6 (42,9) | 2 (40) |
| Presentation/discussion of research projects | 12 (37,5) | 3 (23,1) | 7 (50) | 2 (40) |
| Teaching at the bedside | 20 (62,5) | 6 (46,2) | 10 (71,4) | 4 (80) |
| **EVALUATION TOOLS** | **Total, N (%)** | **Small, N (%)** | **Medium, N (%)** | **Large, N (%)** |
| Direct observation of clinical skills (e.g. diagnostic/therapeutic procedure setting) | 14 (43,8) | 4 (30,8) | 8 (57,1) | 2 (40) |
| Direct observation of procedural skills (e.g. rachicentesis) | 11 (34,4) | 3 (23,1) | 7 (50) | 1 (20) |
| Discussion of errors in diagnosis/treatment | 5 (15,6) | 2 (15,4) | 3 (21,4) | 0 (0) |
| Discussion of specific clinical cases | 10 (31,3) | 5 (38,5) | 4 (28,6) | 1 (20) |
| Feedback from multiple sources (colleagues, nurses and other professionals) | 9 (28,1) | 2 (15,4) | 5 (35,7) | 2 (40) |
| Feedback from patients on the ward and in the outpatient clinic | 4 (12,5) | 2 (15,4) | 1 (7,1) | 1 (20) |
| Frontal, written and/or oral examination | 22 (68,8) | 8 (61,5) | 9 (64,3) | 5 (100) |
| Personal booklet evaluation | 6 (18,8) | 3 (23,1) | 2 (14,3) | 1 (20) |

The data are taken from questions Q18 to Q20.

**Figure 3.1. Theoretical knowledge mean score, total and by school size.**

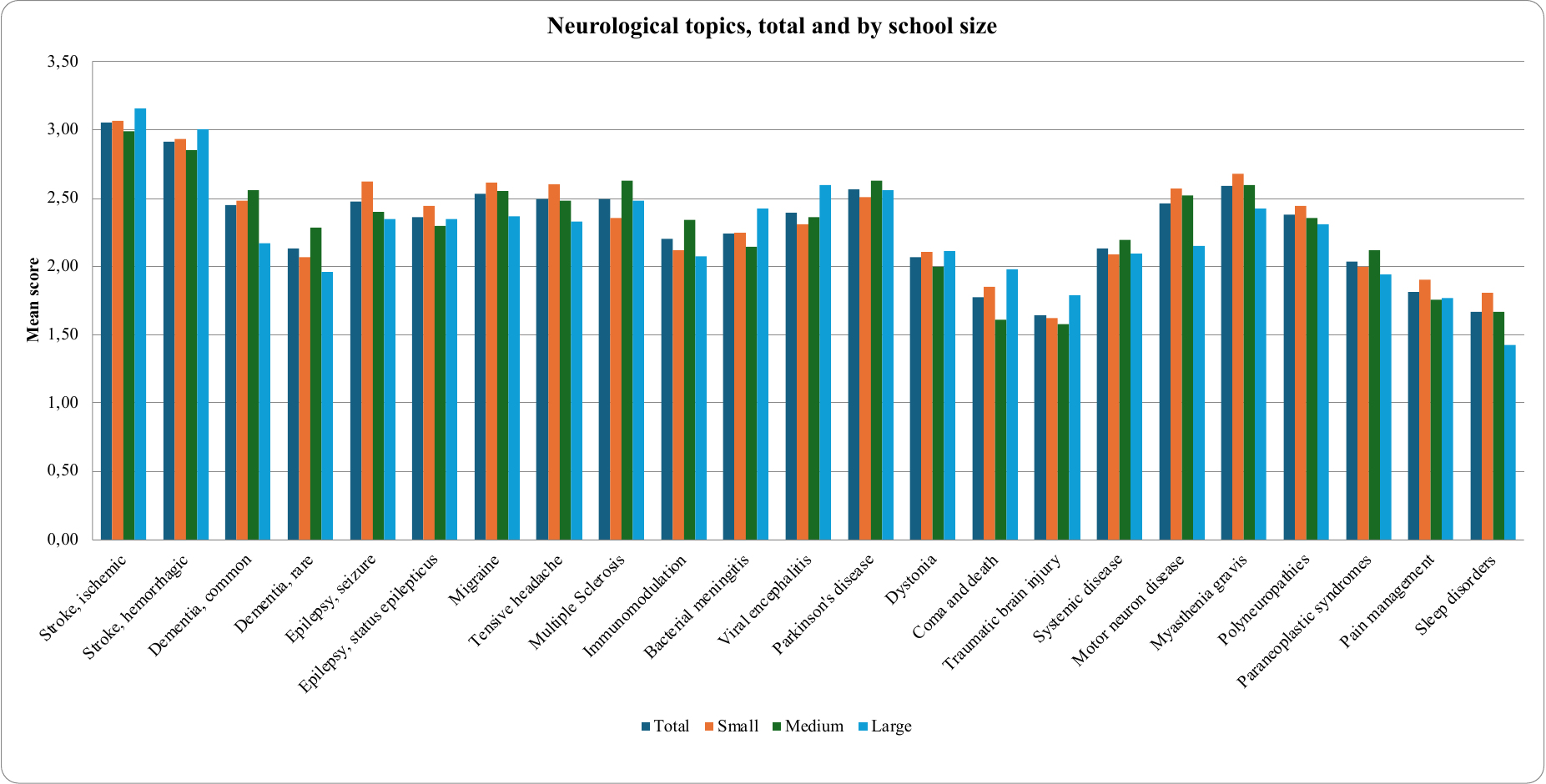

**Figure 3.2. Diagnostic techniques mean score, total and by school size.**

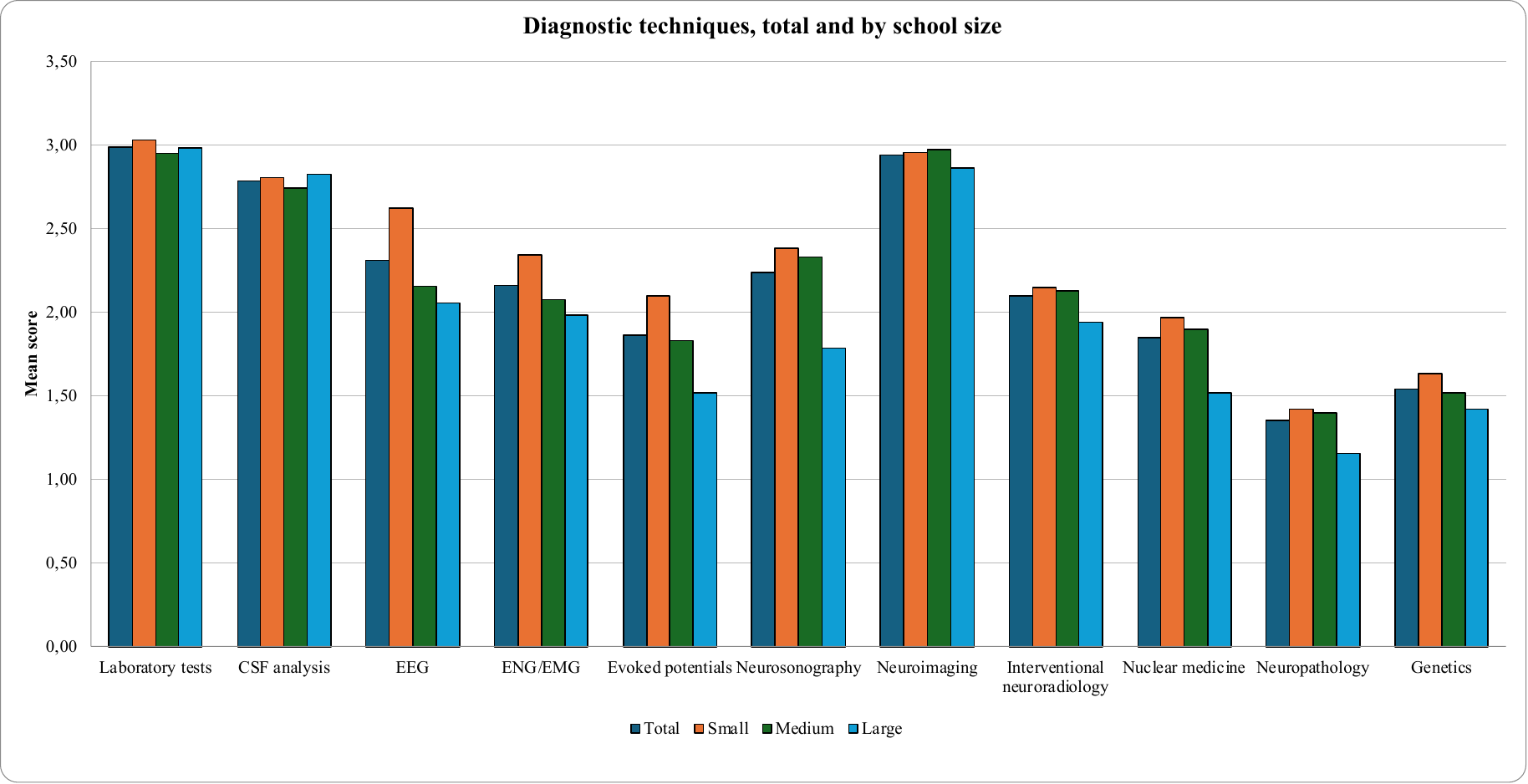

**Figure 3.3. Practical skills mean score, total and by school size.**

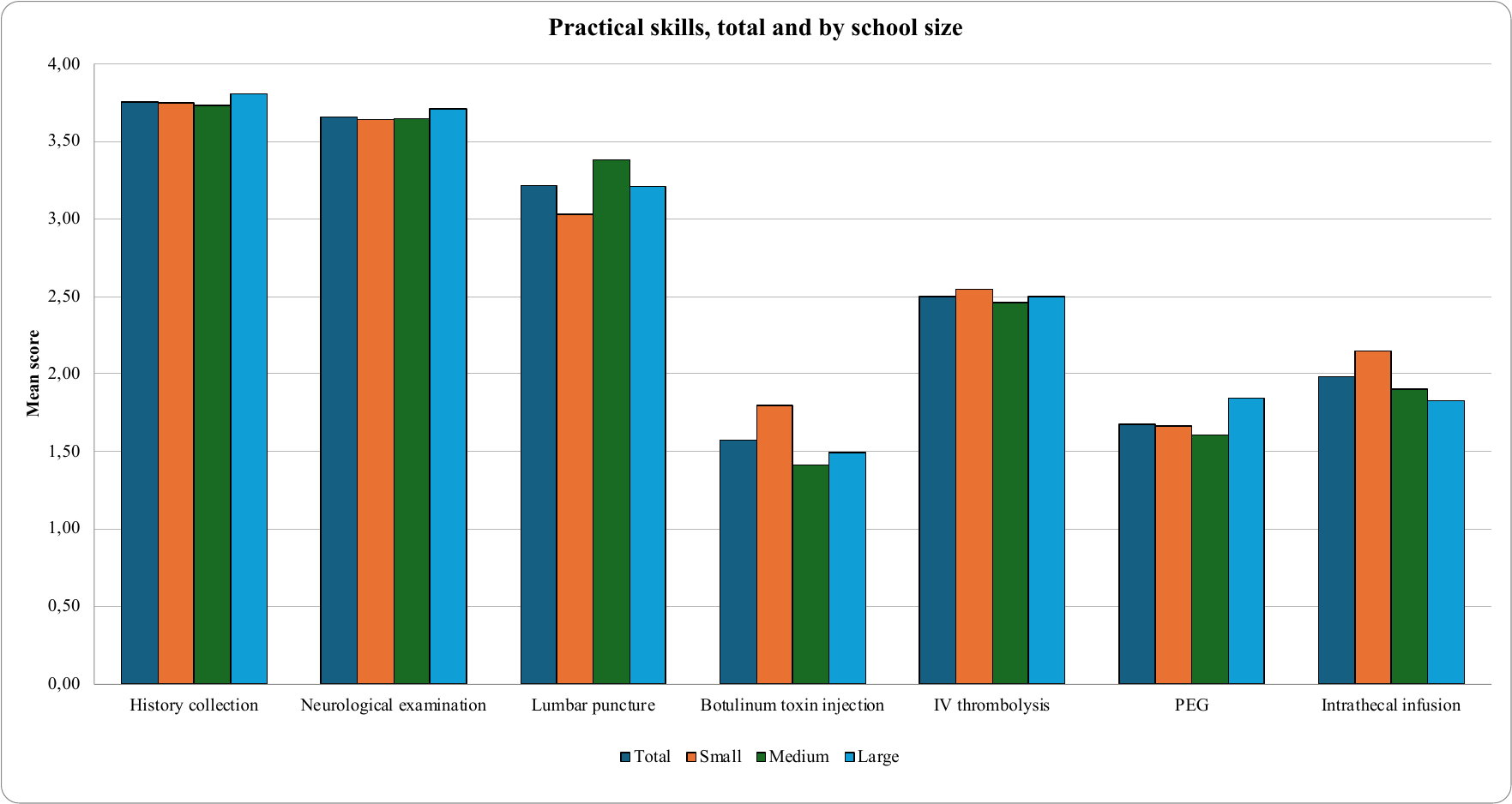
